# Comparative Analysis of Anisotropic Tensile Strength of Human Abdominal Skin

**DOI:** 10.1101/2025.11.21.25340691

**Authors:** Adrian E. Rodrigues, Anmar Abu-Romman, Brigid Coles, Thomas A. Mustoe, Seok Jong Hong, Robert D. Galiano

**Author notes:** **Correspondence:** Dr. Seok Jong Hong, Northwestern University, Feinberg School of Medicine, Division of Plastic Surgery, 300 E. Superior St., Tarry 4-720, Chicago, IL, 60611, USA., Dr. Robert D. Galiano, Northwestern University, Feinberg School of Medicine, Division of Plastic Surgery, Lavin Pavilion, Floor 20, Suite 2060, 259 East Erie Street, Chicago, IL 60611, USA. **Source of Funding:** No funding was received for this research. **Ethics approval statement:** Human specimens were obtained from consented patients undergoing elective abdominoplasties under an Institutional Review Board approved study.

## Abstract

The mechanical properties of human skin have been investigated for decades, yet studies using fresh human tissue remain limited. In this study, we examined the anisotropic behavior of human abdominal skin by assessing tensile strength in panniculectomy specimens obtained from eight female patients. Rectangular 7-mm–long skin samples (0.5, 1.0, and 1.5 cm widths) were excised from each pannus specimen and subjected to uniaxial dynamic tensometry, with the long axis oriented either parallel or perpendicular to Langer’s lines. Samples tested parallel to Langer’s lines exhibited a 2- to 3-fold greater maximum load capacity compared with those tested perpendicular. No statistically significant correlation was detected between tensile strength and body mass index (BMI), skin thickness, or age. Histology confirmed that higher tensile strength occurred when dermal collagen fibers were aligned parallel to the direction of the applied load. These findings demonstrate that collagen orientation strongly influences the mechanical behavior of human skin, with implications for surgical planning, implantable prosthetic design, and wound healing.

## INTRODUCTION

The mechanical properties of human skin have long been studied with the utilization of observational science as the primary method applied during its infancy. In its youth, some well-known findings came from scholars such as Guillaume Dupuytren and Karl Langer, but in truth there were many other well-regarded pioneers.^1,2^ Over time during the 20th century, new discoveries with the use of primitive equipment, yet contemporary scientific methods, were uncovered. Skin mechanical testing using excised specimens from cadavers displayed distinctions between age groups, sex, anatomical location, and even the directionality of the load applied — a property now commonly described as anisotropy.^3-6^ Although these were early adopted approaches, metrics such as elongation, stress-strain curves, and viscoelastic properties were successfully captured. Later, with the introduction of spring and suction devices, a newer in vivo skin parameter could be assessed with Young’s modulus, thereby providing a means to evaluate skin pliability and stiffness by measuring skin stress and strain.^7,8^ Now, with modern tensile equipment and high-speed cameras, excised tissue can undergo quasi-static, cyclic, and high-speed dynamic testing, with some instruments capable of reaching speeds that replicate the effect of high-impact trauma.^9^

Undoubtedly, the mechanical properties of skin have a good history, but testing on fresh human tissue has been limited; most literature cites the use of cadaver or animal specimens.^10-14^ In this study, we look further into the anisotropic behavior of human skin by analyzing its tensile strength to determine its maximum load. By pulling skin of different widths — either parallel to the collagen network known as Langer’s lines or perpendicular to it — we gained a better understanding of the diverging strengths that are consequent to its anisotropic property. By using human pannus samples obtained post-panniculectomy from patients undergoing abdominoplasties, the data closely aligns with the likely behavior seen in intact, healthy living skin. This is important because tissue begins to weaken once blood perfusion is lost through processes such as autolysis and enzymatic degradation, and the length of time since death in a corpse, or time from death until a body is preserved, is commonly not known in cadaveric tissues. Accordingly, this study sought to refine existing knowledge and support therapeutic applications that depend on the mechanical properties of human skin that most closely resemble living tissue.

## METHODS

### Tensile strength measurement

After successful panniculectomy from patients undergoing abdominoplasty, excised pannus specimens of the lower abdomen were immediately transferred out of the surgical suite and stored at 4°C overnight prior to testing. On the following day, rectangular mappings were drawn on the skin of the pannus with the following three dimensions: 0.5 cm × 7.0 cm, 1.0 cm × 7.0 cm, and 1.5 cm × 7.0 cm, with some of the mappings oriented vertically (perpendicular to Langer’s lines) and others oriented horizontally (parallel to Langer’s lines), with collagen lines referenced from Cox (1941).^15^ Skin rectangular mappings were then excised from each pannus, producing rectangular specimens with intact epidermal and dermal layers. The samples were then subjected to uniaxial dynamic tensometry along their 7.0 cm long axis, testing maximum load captured in Newtons (N) with an Instron model 5542 (Instron Corporation, Canton, Massachusetts, USA) equipped with a 500 N load cell set at a crosshead speed of 50 mm/minute (**Figure 1**). All skin specimens that were tested display fatigue at the center of the specimen and not on surfaces adjacent to the grips holding the skin. Eight female patients with a BMI (body mass index) indicating either overweight or obese habitus were included in the study. The number of skin specimens tested from each corresponding pannus is also noted (**Table 1**).

**Table 1.** Patient body mass index (BMI) and age presented in decade-based categories, along with skin specimen count obtained from each patient. (H = horizontal, V = vertical). Specimen count varies across patients due to skin injury (electrosurgery), presence of scars, and/or pannus morphology.

| BMI | Age | 0.5 cm<br>Sample<br>Count | 1.0 cm<br>Sample<br>Count | 1.5 cm<br>Sample<br>Count |
| --- | --- | --- | --- | --- |
| 26 | In their 50's | H=0, V=2 | H=3, V=5 | H=0, V=5 |
| 26 | In their 50's | H=3, V=2 | H=3, V=3 | H=0, V=2 |
| 27 | In their 30's | H=0, V=3 | H=3, V=11 | H=0, V=3 |
| 30 | In their 40's | H=0, V=0 | H=0, V=3 | H=0, V=0 |
| 30 | In their 50's | H=0, V=1 | H=3, V=4 | H=0, V=1 |
| 31 | In their 50's | H=0, V=2 | H=7, V=8 | H=0, V=2 |
| 32 | In their 40's | H=0, V=0 | H=2, V=6 | H=0, V=0 |
| 33 | In their 50's | H=0, V=3 | H=5, V=9 | H=0, V=3 |

**Figure 1.**
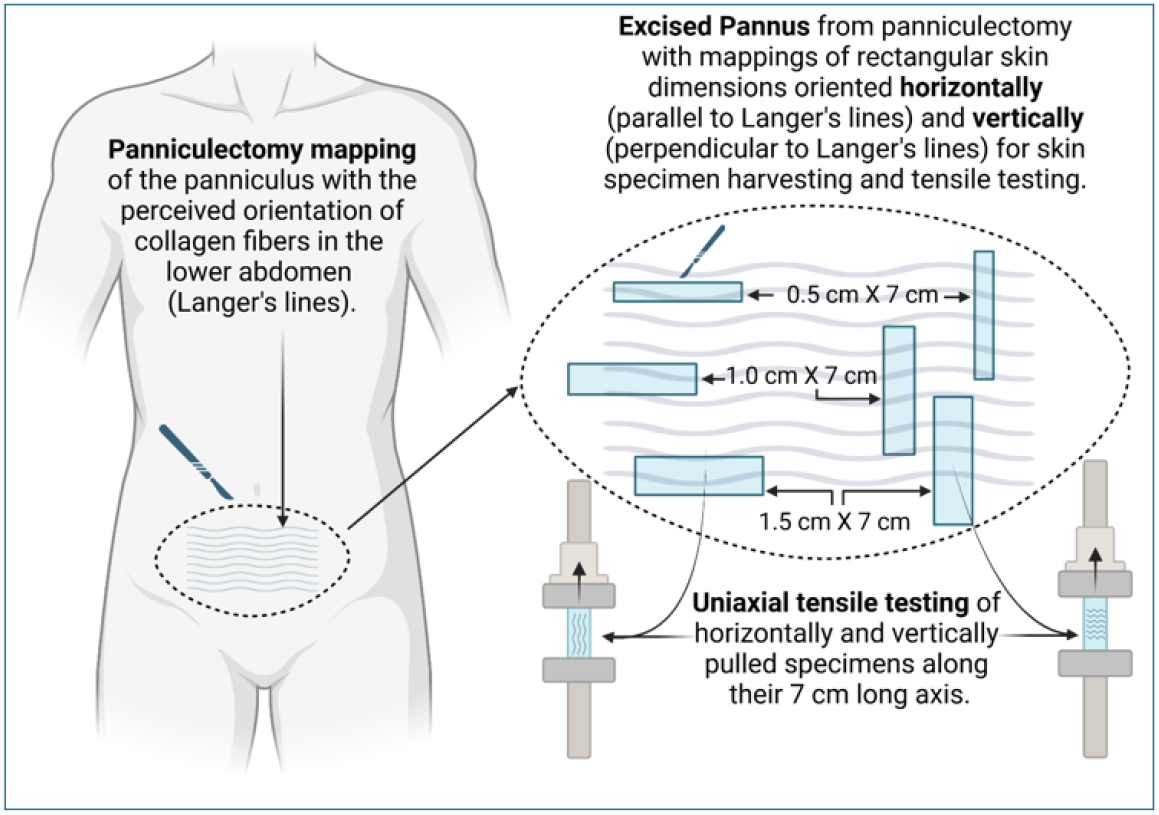
Schematic representation of pannus location, skin specimen mapping and related tensometry.

### Histological analysis

Some rectangular skin samples were solely excised for histological analysis. These were extracted from panni and fixed in 10% neutral-buffered formalin, serially dehydrated, embedded in paraffin and cut to 5μm-thick sections. Samples were then deparaffinized and stained with Masson’s trichrome or hematoxylin and eosin (H&E) according to standard protocols. Skin thickness quantification included epidermal and dermal layers, measured in millimeters (mm). Images were obtained with a Nikon Eclipse 50i microscope and Nikon NIS Elements software (Nikon Instruments, Melville, NY).

### Statistical analysis

Statistical analyses were performed using Prism 9 (GraphPad, San Diego, CA). Statistics are represented as mean ± standard deviation, n = number of independent rectangular skin specimens excised for testing, *P < 0.05, **P < 0.01, ***P < 0.001, ****P < 0.0001, and not significant (ns) P > 0.05. Bar graphs show data assessed with a two-tailed unpaired t-test in pairwise comparisons and an ordinary one-way ANOVA in multiple comparisons. Scatter plots show a Pearson correlation between the quantified variables.

## RESULTS

Specimens pulled parallel to collagen orientation (horizontally pulled) displayed a significant difference between 0.5 cm and 1.0 cm widths tested. Doubling the width from 0.5 cm to 1.0 cm produced a more than double increase in maximum load, making them highly significant. All of the wider 1.5 cm X 7.0 cm samples (5 in total) that were pulled horizontally exceeded the tensometer’s load cell capacity of 500 N, and therefore data from samples 1.5 cm wide were not captured but can be inferred to exceed 500 N, nevertheless given that they resisted at least 500 N demonstrates an impactful gain in maximum load (**Figure 2A**).

**Figure 2.**
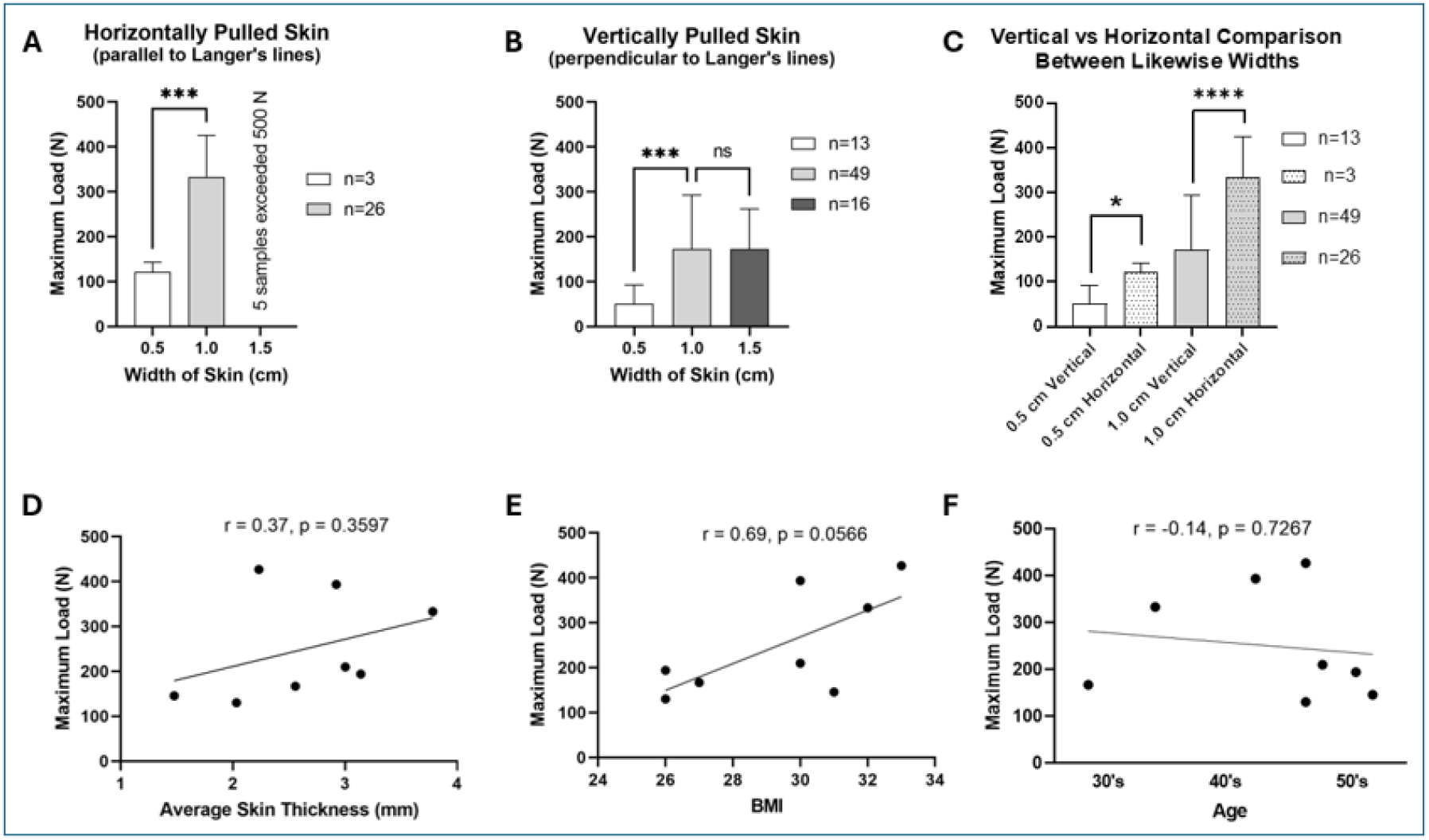
Graphical representation of results **A:** Horizontally pulled skin comparing 0.5 cm and 1.0 cm wide skin samples. The 1.5 cm samples exceed our instrument’s capacity to capture the maximum load. **B**: Vertically pulled skin, showing 0.5 cm, 1.0 cm and 1.5 cm wide samples, showing no gain in load between the latter two widths. **C**: Vertical vs horizontal comparison between likewise widths, showing the stepwise anisotropic characteristics between 0.5 cm and 1.0 cm wide skin samples. **D-F**: Scatter plot showing no correlation between maximum load and the average skin thickness (D), body mass index (BMI) (E), or age (shown as decade-based) (F).

Skin samples perpendicular to collagen (vertically pulled) also showed significant differences in maximum load between 0.5 cm and 1.0 cm wide samples, however increasing the width to 1.5 cm failed to show any increase in maximum load (**Figure 2B**).

To better visualize the anisotropic behavior of the skin specimens, vertical vs horizontal graphical comparisons were made between equal widths. Specimens 0.5 cm wide showed a marginally significant (P = *) difference in maximum load gain when pulled horizontally, whereas 1.0 cm wide samples displayed an extremely significant (P = ****) difference in load gain when pulled horizontally (**Figure 2C**). No correlation could be made between maximum load and skin thickness, BMI or age (**Figure 2D-F**).

Langer’s lines correspond to the orientation of collagen fibers within the dermis. Making incisions along Langer’s lines typically results in less scarring and faster healing due to the natural direction of collagen alignment. The histology of the skin of the lower abdomen supports the existence of distinctive collagen directionality. By fixing skin samples perpendicular and parallel to Langer’s lines, one can visualize the cross-sectional **(Figure 3, A & B)** and longitudinal arrangement **(Figure 3, C & D)** of the collagen bundles in the dermis, respectively, supporting the presence of a predominantly directional network of aligned fibers.

**Figure 3.**
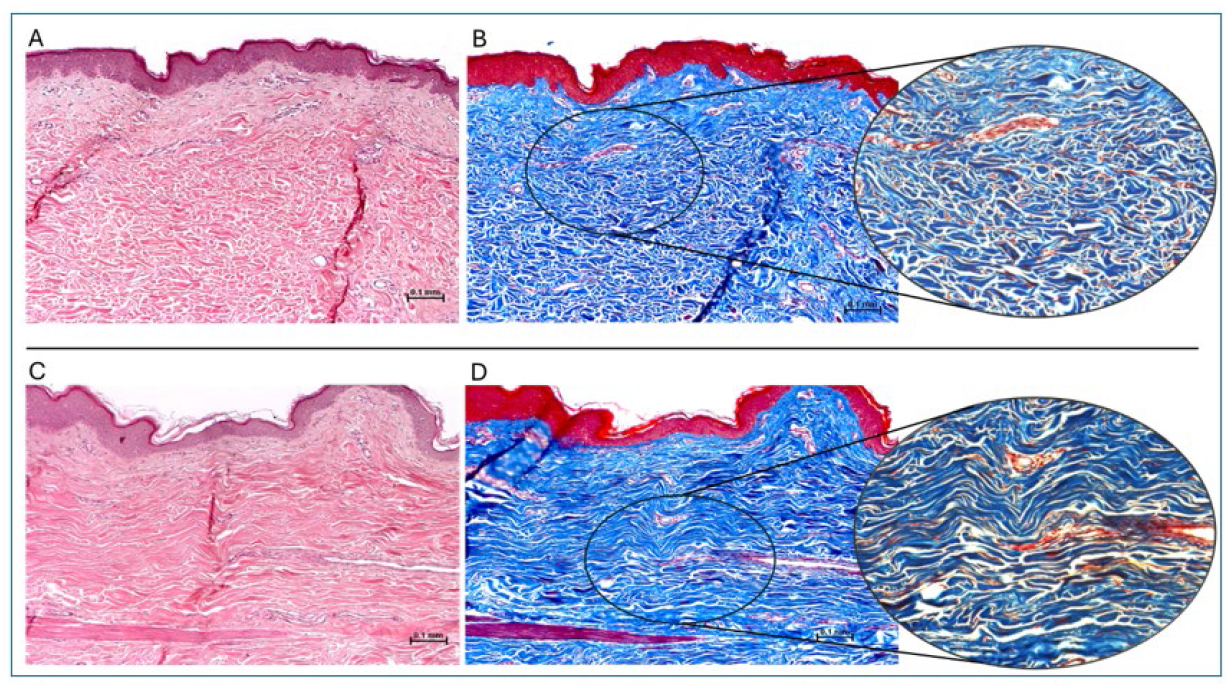
Collagen orientation Skin histology witn H&E (**A**) ano Masson’s trichrome staining (**B**) ot skin aligned | perpendicular to Langer’s lines, showing small fragments of collagen bundles due to perpendicular sectioning of their longitudinal trajectory. H&E (**C**) and Masson’s trichrome staining (**D**) with skin aligned parallel to Langer’s lines showing a dominant elongated direction of collagen bundles.

## DISCUSSION

Materials like rope and wire can best resist fatigue when a load is equally distributed in the direction of fiber alignment because it allocates the burden collectively across all fibers. On the other hand, when a force is applied perpendicular to the alignment of fibers — against their natural orientation — equal distribution of the load burden is lost and therefore collective effort cannot be attained. Thus, this directional tolerance is the mechanical anisotropic property we described earlier, and its presence in biological studies has been supported, likely because human and animal collagen in unwounded skin favors a dominant linear orientation,^10,16^ and the histology from our specimens further support the existence of this alignment.

Thus, in this study, we not only witnessed directional differences influenced by collagen alignment, but also the exponentially diverging anisotropic tolerances when testing different widths in opposing directions. At 0.5 cm in width, the directional differences were approximately 50 N and 100 N respectively, and at 1.0 cm they diverged from approximately 150 N to 300 N (Figure 2C). However, failure to see an increase in 1.5 cm wide samples pulled vertically (Figure 2B), coupled with the fact that 1.5 cm wide samples pulled horizontally topped the 500 N load cell capacity (Figure 2A), supports the exciting impression that the anisotropic mechanics of human skin exponentially diverge with increasing widths. This illustrates that collagen exhibits an exponential gain in load-bearing capacity with subsequently wider specimens when pulled along its orientation, and displays a diminishing return (fails to increase in load capacity with increasing widths) when the force is perpendicular to the collagen alignment. Though, given the non-correlating results, we cannot say that these findings are related to skin thickness, BMI or patient age (Figure 2, D-F).

Building on these findings, it is essential to explore how skin mechanics are influenced by factors such as wounds. For instance, in injured skin that has undergone healing, the restructuring and remodeling of collagen is known to result in unique mechanical properties. Compared to normal skin, hypertrophic scars exhibit decreased strength, reduced extensibility, and increased stiffness.^17^ Some of these differences may be attributed to the erratic reorganization and reduced diameter of collagen fibers in scar tissue, as supported by several authors.^18,19^ Clearly, collagen plays a critical structural role, whether the skin is injured or not. However, this study has several limitations. The investigation was restricted to a single dynamic test without cyclic or quasi-static analysis. Moreover, the analysis focused exclusively on abdominal skin, which may not be representative of other body regions. Also, patient specimen samples were limited to obese and overweight females, potentially excluding variations in normal-weight individuals or males. Finally, the load cell capacity of the Instron device restricted testing for 1.5 cm-wide skin samples pulled horizontally. Despite these limitations, further studies on skin mechanics are necessary, especially human skin that has been subjected to tissue expansion devices. Such research will deepen our understanding of the strengths and limitations of skin mechanics, which will lend importance to clinical settings.

## CONCLUSION

This study reveals the remarkable anisotropic mechanical properties of human skin, demonstrating how its load-bearing capacity critically depends on collagen fiber orientation. By testing fresh panniculectomy specimens, we observed that skin pulled parallel to collagen lines exhibits exponentially increasing strength with specimen width, while perpendicular loading shows diminishing mechanical resistance. These findings highlight the complex biomechanical nature of skin and provide valuable insights for medical applications such as implantable prosthetics, wound closure, and surgical mapping. While our research is limited by sample demographics and testing conditions, it underscores the importance of studying human tissue to understand skin’s mechanical behavior. Future investigations should explore diverse populations, anatomical regions, and various testing methods to further elucidate skin’s intricate mechanical properties.

## Data Availability

All data produced in the present work are contained in the manuscript

## Acknowledgement

Some figures were created with Biorender.com

